# Early and Midterm Outcomes of Transcatheter Aortic-Valve Replacement with Balloon-Expandable Versus Self-Expanding Valves: A Systematic Review and Meta-Analysis

**DOI:** 10.1101/2020.06.20.20136143

**Authors:** Xin-Lin Zhang, Xiao-Wen Zhang, Zhong-Hai Wei, Li-Na Kang, Rong-Fang Lan, Jian-Zhou Chen, Jun Xie, Lian Wang, Wei Xu, Biao Xu

## Abstract

**Background:** The comparative performances of transcatheter aortic-valve replacement (TAVR) with balloon-expandable valves (BEV) and self-expanding valves (SEV) in severe aortic stenosis remain unclear.

**Purpose:** To compare the early (30-day) and midterm (1-year) mortality and cardiovascular outcomes of BEV with SEV.

**Data Sources:** PubMed, EMBASE, and the Cochrane Library from inception until February 13, 2020.

**Study Selection:** 3 randomized controlled trials (RCTs) and 12 propensity-score matched (PSM) studies, with 37,958 patients.

**Data Extraction:** 2 reviewers independently extracted study data and rated study quality. **Data Synthesis:** Compared with SEV, BEV was associated with significantly lower mortality at 30 days (OR 0.77, 95% CI 0.71–0.83, *P<*0.00001, *I*^*2*^=0) and a trend toward lower mortality at 1 year (OR 0.88, 95% CI 0.78–1.00, *P=*0.05, *I*^*2*^=15.8%), mainly driven from PSM studies, but regardless of valve generations and SEV types. 30-day and 1-year cardiovascular mortality, 30-day incidences of moderate to severe paravavular leak, procedural contrast agent volume and procedure time were lower, but transvalvular pressure gradient was higher in BEV than SEV. 30-day incidences of permanent pacemaker implantation (PPI), acute kidney injury, stroke, major bleeding, major vascular complications and rehospitalization were not statistically different between BEV and SEV. Early-generation SEV was associated with a higher 30-day PPI risk than corresponding BEV comparators. PPI risk was lower in ACURATE neo but higher in Evolut R SEV, both compared with SAPIEN 3 BEV.

**Limitations:** Study-level but not patient-level data; residual confounders in PSM studies; study designs and patient characteristics were heterogeneous.

**Conclusions:** Compared with SEV, BEV might be associated with lower early and midterm mortality. Results from adequately powered RCTs with long-term follow-up are critically needed to confirm these findings.

**Registration:** PROSPERO (CRD42020172889).

**Funding Source:** National Natural Science Foundation of China (NO. 81600312).

## Introduction

Transcatheter aortic-valve replacement (TAVR) has emerged as a safe and effective therapy for patients with severe aortic stenosis at all-range of surgical risks. A number of trials have shown noninferior or superior performance of TAVR over surgical aortic-valve replacement (SAVR) (1). TAVR has overtaken SAVR as the most common approach for aortic valve replacement in several countries (2).

Among all TAVR valve systems, the balloon-expandable and self-expanding systems gained the most amount of data. Current guidelines provide recommendation for TAVR without emphasis on valve systems (3, 4), but it’s important to compare different valve systems as TAVR indications expand to younger, lower-risk patients with longer life expectancy. A network meta-analysis (5) showed no significant difference in short-term mortality between balloon-expandable valves (BEV) and self-expanding valves (SEV), mainly driven from indirect comparisons and with considerable heterogeneity. Direct comparisons of BEV with SEV have been lacking until recent publications of several small-to-moderate-sized randomized controlled trials (RCTs) (6-8). All these trials were underpowered to detect rare outcomes such as mortality. A number of larger-scale propensity-score matched (PSM) studies (9-11) were recently published and might provide complementary insights for RCTs. In this context, we performed a meta-analysis of head-to-head RCTs and PSM studies to determine the early and midterm performances of TAVR with BEV versus SEV.

## Methods

We reported the meta-analysis following the Preferred Reporting Items for Systematic Reviews and Meta-Analyses (PRISMA) guideline (12).

### Data Sources and Searches

PubMed, the Cochrane Central Register of Controlled Trials, and EMBASE were systematically searched through February 13, 2020 (X.L.Z. and X.W.Z.). The computer-based searches, reviewed by a medical librarian, combined terms and combinations of keywords which included self-expanding, balloon-expandable, and transcatheter aortic-valve. Two investigators (X.L.Z. and X.W.Z.) also independently reviewed the reference lists of identified studies and relevant reviews to search for other relevant studies.

### Study Selection

Two reviewers (X.L.Z. and X.W.Z.) independently screened the titles and abstracts for eligibility, and retrieved full-text for those with potential relevance. Conflicts were handled by consensus, or resolved by a third investigator (L.K.) if necessary. Eligible studies (1) evaluated patients with severe aortic stenosis who received a TAVR therapy, (2) compared BEV with SEV, (3) reported at least one outcome of interest, (4) were RCTs or PSM studies. We excluded adjusted observational studies without PSM, observational studies without adjustment, single-arm studies, studies performed in patients with degenerated aortic surgical bioprostheses, and studies comparing devices other than SEV and BEV.

### Outcome Measures

The primary outcomes were early (30-day) and midterm (1-year) all-cause mortality. Secondary outcomes included 30-day and 1-year cardiovascular mortality, 30-day stroke, permanent pacemaker implantation (PPI), major bleeding, major vascular complications (MVC), acute kidney injury (AKI), rehospitalization, and moderate to severe paravavular leak (PVL). We also included procedure outcomes such as transvalvular pressure gradient, contrast agent volume, procedure time, and fluoroscopy time.

### Data Extraction and Quality Assessment

Two investigators (X.L.Z. and X.W.Z.) independently extracted study data using a prespecified form, evaluated the risk of bias of RCTs using the Cochrane Collaboration’s risk of bias tool (13) and the quality of PSM studies using the Newcastle–Ottawa Scale (14). We also evaluated the quality and appropriateness of PSM methods used in the observational studies (15).

### Data Synthesis and Statistical Analysis

Summary measures are presented as odds ratios (ORs) for binary outcomes and mean differences for continuous outcomes, and pooled using random-effect models (DerSimonian–Laird method) with Hartung-Knapp-Sidik-Jonkman variance correction (16). Heterogeneity was assessed using the Q and *I*^*2*^ statistics. Stratified analyses were performed according to study designs (RCTs and PSM studies), valve generations (early-generation and new-generation) and SEV types (Boston Scientific ACURATE neo, Medtronic Evolut R, Medtronic CoreValve, and Abbott Portico). Between-subgroup differences were evaluated with the Q-test for heterogeneity. We performed random-effects meta-regression analysis to outline the association between early all-cause mortality and other potentially relevant outcomes, including moderate to severe PVL, PPI, and stroke. Potential publication bias was examined by visual inspection of the asymmetry in funnel plots. Both the studies of Deharo (11) and Van Belle (9) originated from national registries of French; we extracted data before last quarter 2014 in the study of Van Belle to avoid overlap with the study of Deharo. All meta-analyses were conducted with the R statistical programming environment, version 4.0.0 and the Review Manager, version 5.3, and a 2-tailed *P* value<0.05 was considered statistically significant.

## Results

### Study selection and characteristics

We included 15 studies—3 RCTs (6-8) and 12 PSM studies (9-11, 17-25)—with 37,958 patients (19,053 with BEV and 18,905 with SEV) (Figure 1). The mean age was 82.6 years and 42.4% were male. When reported, the mean Logistic EuroSCORE ranged from 14 to 22.8, the mean STS risk score ranged from 3.7 to 9.3. Most procedures were performed through transfemoral approach. Baseline characteristics were presented in Appendix Table 1 to 4. All trials were deemed as having low risk of bias (Appendix Table 5). The nature of the intervention made trials blinded for clinicians impossible. All PSM studies achieved high scores on the Newcastle–Ottawa Scale, ranging from 8 to 9 stars (Appendix Table 6). The quality of reporting on PSM varied (Appendix Table 7), detailed in the discussion section.

**Figure 1.**
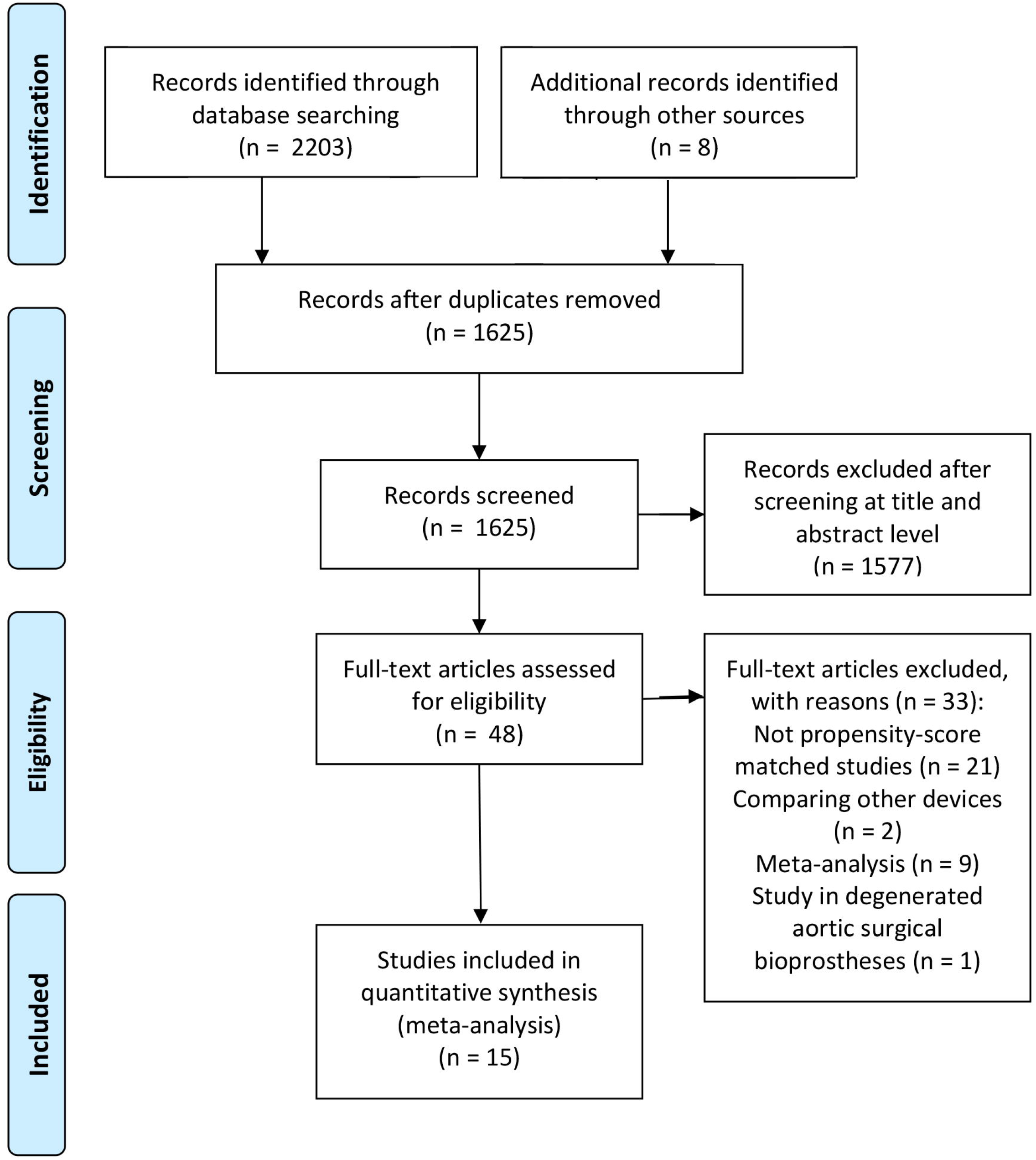
PRISMA flow diagram.

### Early and midterm all-cause mortality

Overall, BEV was associated with a significantly lower 30-day all-cause mortality compared with SEV (OR 0.77, 95% CI 0.71–0.83, *P<*0.00001, *I*^*2*^=0) (Figure 2A). No publication bias was observed (Appendix Figure 1). Stratified analysis showed a nonstatistically difference in RCTs (OR 0.60, 95% CI 0.19–1.90, *P=*0.19, *I*^*2*^=0) and a statistically significant risk reduction in PSM studies (OR, 0.76, 95% CI, 0.67–0.85, *P<*0.001, *I*^*2*^=0), without significant difference between subgroups (*P* value for interaction=0.39) (Table 1). 30-day all-cause mortality was significantly lower in BEV regardless of valve generations (early-generation: OR 0.74, 95% CI 0.65–0.85, *P=*0.002, *I*^*2*^=0; new-generation: OR, 0.77, 95% CI, 0.68–0.87, *P=*0.001, *I*^*2*^=0) (Figure 2B, Table 2). Because all BEV were all of the Edwards SAPIEN family, but SEV were from 3 different companies, we performed subgroup analysis based on SEV types, and found consistently lower 30-day all-cause mortality in BEV (SAPIEN 3 vs. Evolut R: OR 0.78, 95% CI 0.75–0.82, *P=*0.002, *I*^*2*^=0; SAPIEN/SAPIEN XT vs. CoreValve: OR 0.74, 95% CI 0.65–0.85, *P=*0.002, *I*^*2*^=0) (Appendix Figure 2). Comparison between ACURATE neo, Portico and SAPIEN 3 valves seemed underpowered (SAPIEN 3 vs. ACURATE neo: OR 0.56, 95% CI 0.28–1.12, *P=*0.08, *I*^*2*^=0).

**Table 1.** Risk estimates of 30-day and 1-year outcomes for TAVR with balloon-expandable versus self-expanding valves according to study designs.

| Outcomes | Randomized controlled trials |  |  |  | Propensity-score matched studies |  |  |  |  | Overall |  |  |  |
| --- | --- | --- | --- | --- | --- | --- | --- | --- | --- | --- | --- | --- | --- |
| | No of | OR (95% CI) | $I^2$ | $P$ value | No of | OR (95% CI) | $I^2$ | $P$ value | $P$ value for | No of | OR (95% CI) | $I^2$ | $P$ value |
|  | studies |  | (%) |  | studies |  | (%) |  | interaction* | studies |  | (%) |  |
|  | (patients) |  |  |  | (patients) |  |  |  |  | (patients) |  |  |  |
| 30-day follow-up |  |  |  |  |  |  |  |  |  |  |  |  |  |
| All-cause mortality | 3 (1418) | 0.60 (0.19–1.90) | 0 | 0.19 | 11 (36540) | 0.76 (0.67–0.85) | 0 | <0.001 | 0.39 | 14 (37958) | 0.77 (0.71–0.83) | 0 | <0.00001 |
| Cardiovascular mortality | 2 (980) | 0.62 (0.24–1.62) | 6.8 | 0.33 | 5 (22336) | 0.77 (0.60–1.00) | 0 | 0.05 | 0.67 | 7 (23316) | 0.77 (0.64–0.93) | 0 | 0.01 |
| PPI | 3 (1418) | 0.69 (0.24–1.99) | 57.4 | 0.27 | 10 (36052) | 0.77 (0.46–1.27) | 96.7 | 0.27 | 0.75 | 13 (37470) | 0.74 (0.50–1.09) | 95.1 | 0.12 |
| Moderate to severe PVL | 3 (1418) | 0.28 (0.12–0.65) | 0 | 0.02 | 9 (7286) | 0.44 (0.30–0.64) | 55.4 | < 0.0001 | 0.08 | 12 (8704) | 0.40 (0.29, 0.55) | 47.5 | <0.00001 |
| AKI | 3 (1418) | 0.56 (0.11–2.75) | 45.6 | 0.26 | 5 (1773) | 0.85 (0.45–1.60) | 0 | 0.51 | 0.33 | 8 (3191) | 0.75 (0.48–1.16) | 0 | 0.16 |
| Stroke | 3 (1418) | 2.30 (0.36–14.7) | 2.1 | 0.19 | 10 (15622) | 0.73 (0.59–0.90) | 0 | 0.01 | 0.01 | 13 (17040) | 0.89 (0.62–1.27) | 21.9 | 0.48 |
| MVC | 3 (1418) | 0.86 (0.39–1.93) | 0 | 0.51 | 8 (6058) | 1.10 (0.87–1.38) | 0 | 0.38 | 0.26 | 11 (7476) | 1.04 (0.84–1.28) | 5.3 | 0.71 |
| Major bleeding | 3 (1418) | 1.09 (0.35–3.39) | 18.7 | 0.78 | 11 (15622) | 1.08 (0.94–1.24) | 3.4 | 0.24 | 0.98 | 14 (17040) | 1.08 (0.95–1.22) | 2.3 | 0.20 |
| Rehospitalization | 2 (980) | 0.39 (0.05, 3.05) | 46.9 | 0.37 | 3 (22040) | 1.01 (0.59, 1.74) | 53.8 | 0.95 | 0.37 | 5 (23020) | 0.98 (0.67, 1.43) | 33.4 | 0.87 |
| 1-year follow-up |  |  |  |  |  |  |  |  |  |  |  |  |  |
| All-cause mortality | 1 (241) | 1.47 (0.72–3.01) | NA | 0.29 | 7 (26950) | 0.87 (0.77–0.99) | 20.4 | 0.04 | 0.16 | 8 (27191) | 0.88 (0.78–1.00) | 15.8 | 0.05 |
| Cardiovascular mortality | 1 (241) | 1.40 (0.62–3.19) | NA | 0.42 | 3 (25250) | 0.87 (0.79–0.97) | 0 | 0.01 | 0.24 | 4 (25491) | 0.86 (0.77–0.95) | 0 | 0.02 |

AKI, acute kidney injury; MVC, major vascular complication; OR, odds ratio; PPI, permanent pacemaker implantation; PVL, paravavular leak; TAVR, transcatheter
aortic-valve replacement.

**Table 2.** Risk estimates of 30-day outcomes for TAVR with balloon-expandable versus self-expanding valves according to valve generations.

| Outcomes | Early-generation valves |  |  |  | New-generation valves |  |  |  |  |
| --- | --- | --- | --- | --- | --- | --- | --- | --- | --- |
|  | No of studies | OR (95% CI) | *I*<sup>2</sup> | *P* value | No of studies | OR (95% CI) | *I*<sup>2</sup> | *P* value | *P* value for |
|  | (patients) |  | (%) |  | (patients) |  | (%) |  | interaction |
| All-cause mortality | 6 (10855) | 0.74 (0.65–0.85) | 0 | 0.002 | 10 (26612) | 0.77 (0.68–0.87) | 0 | 0.001 | 0.67 |
| PPI | 4 (10367) | 0.31 (0.22–0.44) | 63.0 | 0.002 | 10 (26612) | 0.97 (0.69–1.35) | 84.2 | 0.82 | <0.0001 |
| Moderate to severe PVL | 5 (5067) | 0.47 (0.21–1.05) | 70.3 | 0.06 | 7 (3337) | 0.32 (0.23–0.44) | 0 | 0.0001 | 0.21 |
| AKI | 3 (729) | 0.53 (0.13–2.24) | 0 | 0.2 | 5 (2462) | 0.85 (0.46–1.56) | 0 | 0.49 | 0.24 |
| Stroke | 6 (10855) | 0.76 (0.54–1.07) | 0 | 0.1 | 8 (4694) | 1.00 (0.45–2.22) | 53.9 | 1.0 | 0.45 |
| MVC | 4 (4653) | 1.14 (0.91–1.43) | 0 | 0.17 | 7 (2823) | 0.89 (0.60–1.32) | 0 | 0.48 | 0.15 |
| Major bleeding | 6 (10855) | 0.99 (0.83–1.17) | 0 | 0.86 | 9 (5694) | 1.13 (0.72–1.80) | 37.5 | 0.55 | 0.51 |

2 Early-generation valves included Edwards Sapien/Sapien XT and Medtronic CoreValve, new-generation valves included Edwards SAPIEN3, Medtronic Evolut R, Boston
3 Scientifics Accurate Neo, and Abbott Portico. AKI, acute kidney injury; MVC, major vascular complication; OR, odds ratio; PPI, permanent pacemaker implantation; PVL,
1 paravavular leak; TAVR, transcatheter aortic-valve replacement.

**Figure 2.**
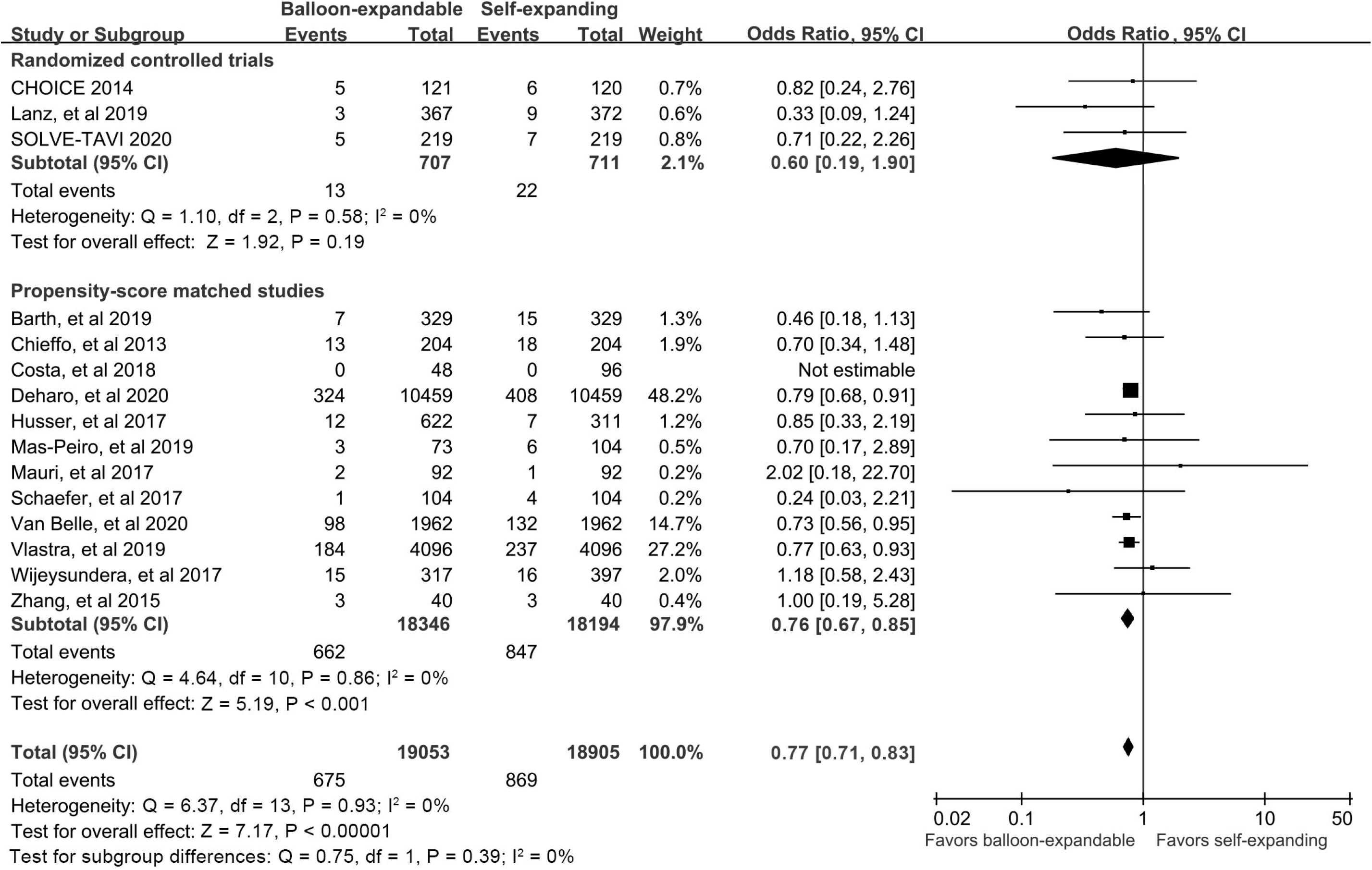
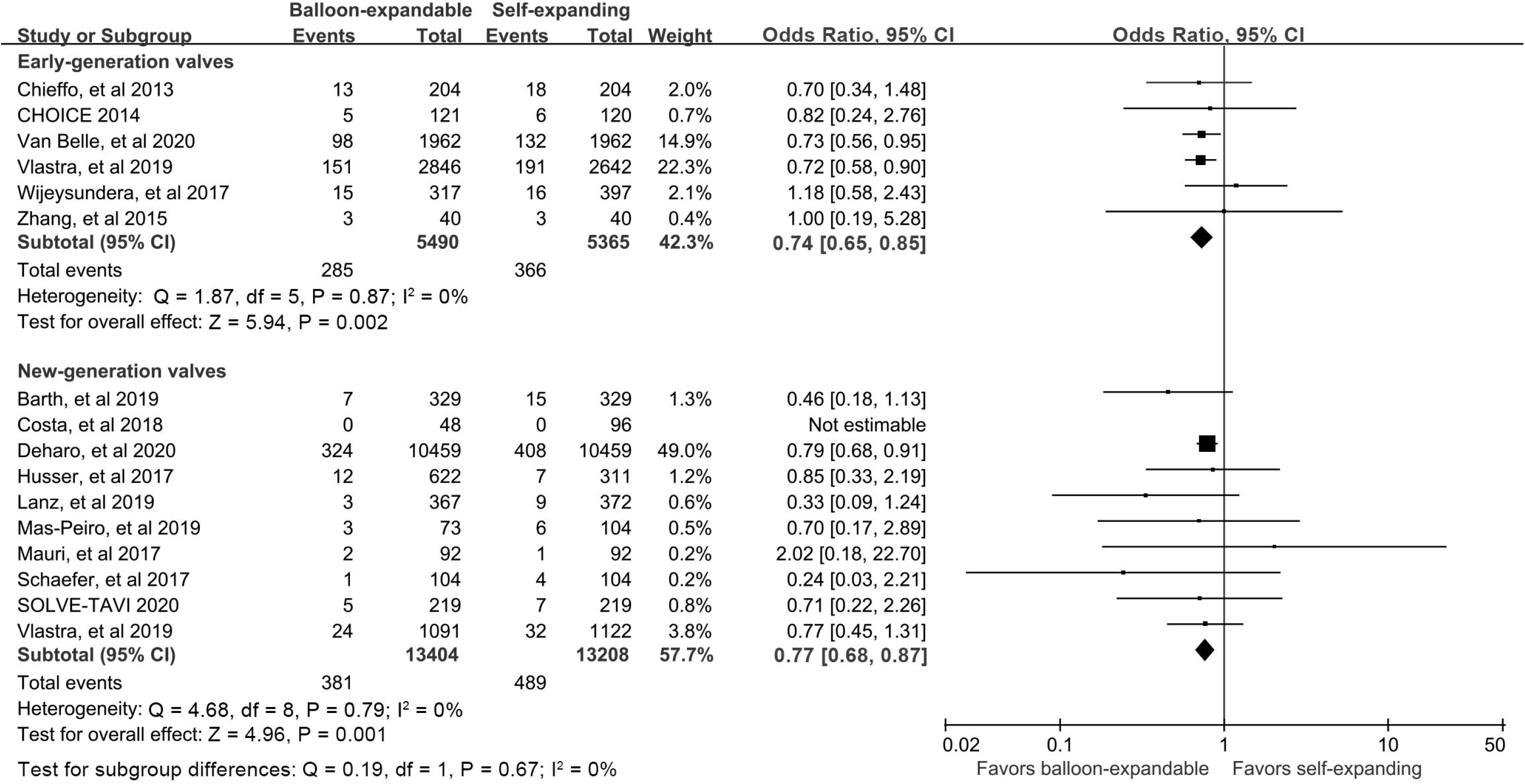
Risk estimates of 30-day all-cause mortality for TAVR with balloon-expandable versus self-expanding valves according to study designs (A) and generations of valves (B).

BEV was associated with a trend toward lower 1-year mortality compared with SEV (OR, 0.88, 95% CI, 0.78–1.00, *P=*0.05, *I*^*2*^=15.8%) (Appendix Figure 3). Only one underpowered RCT reported 1-year mortality (OR, 1.47, 95% CI, 0.72–3.01, *P=*0.29). PSM studies showed a significantly lower 1-year mortality in BEV (OR, 0.87, 95% CI, 0.77–0.99, *P=*0.04, *I*^*2*^=20.4%). No significant difference was detected between subgroups (*P* value for interaction=0.16).

### Early and midterm cardiovascular mortality

The overall incidences of 30-day (OR, 0.77, 95% CI, 0.64–0.93, *P=*0.01, *I*^*2*^=0) and 1-year (OR, 0.86, 95% CI, 0.77–0.95, *P=*0.02, *I*^*2*^=0) cardiovascular mortality were significantly lower in BEV than SEV (Appendix Figure 4 and 5). Stratified analysis showed a statistically significant risk reduction with BEV in PSM studies, but not in RCTs. No significant difference was detected between subgroups.

### Early PPI, moderate to severe PVL

30-day incidence of PPI was not statistically different between BEV and SEV (OR, 0.74, 95% CI, 0.50–1.09, *P=*0.12, *I*^*2*^=95.1%) (Appendix Figure 6). BEV was associated with a significantly lower PPI incidence in early-generation (OR, 0.31, 95% CI, 0.22–0.44, *P=*0.002, *I*^*2*^=63.0%) but not new-generation valves (OR, 0.97, 95% CI, 0.69–1.35, *P=*0.82, *I*^*2*^=84.2%) (Appendix Figure 7). Compared with SAPIEN 3 BEV, PPI incidence was lower in ACURATE neo SEV (OR, 1.45, 95% CI, 1.05–2.01, *P=*0.03, *I*^*2*^=18.4%), but higher in Evolut R SEV (OR, 0.62, 95% CI, 0.39–1.00, *P=*0.05, *I*^*2*^=76.2%), showing significant difference between subgroups (Appendix Figure 8).

30-day incidence of moderate to severe PVL was significantly lower in BEV (OR, 0.40, 95% CI, 0.29–0.55, *P<*0.00001, *I*^*2*^=47.5%), both in RCTs and PSM studies (Appendix Figure 9), early and new-generation valves (Appendix Figure 10 and 11), without significant difference between subgroups.

### Early stroke, AKI, MVC, major bleeding, and rehospitalization

30-day incidence of stroke was not different between BEV and SEV. Stroke incidence was lower in BEV in PSM studies but not RCTs, with significant difference between subgroups (Appendix Figure 12). Analyses based on valve generations and SEV types showed no significant differences (Appendix Figure 13 and 14). There was no significant difference in 30-day AKI, MVC, major bleeding and rehospitalization between BEV and SEV, and all stratified analyses showed similar findings (Appendix Figure 15 to 23).

### Transvalvular pressure gradient, procedural contrast agent volume, procedure time and fluoroscopy time

BEV was associated with a significantly higher transvalvular pressure gradient (Appendix Figure 24), lower contrast agent volume (Appendix Figure 25) and procedure time (Appendix Figure 26) compared with SEV, but fluoroscopy time was similar (Appendix Figure 27).

### Meta-regression analysis

Meta-regression analysis showed a trend toward higher 30-day all-cause mortality with higher incidence of moderate to severe PVL (*P=*0.054) (Appendix Figure 28) and lower incidence of PPI (*P=*0.064) (Appendix Figure 29). No association was observed between 30-day all-cause mortality and risk for stroke (*P=*0.29) (Appendix Figure 30).

## Discussion

In our meta-analysis, BEV might be associated with lower incidences of early and midterm all-cause mortality, though these differences did not reach statistical significance in RCTs. These differences seemed to be independent of valve generations and SEV types, and might be associated with moderate to severe PVL risk differences. 30-day and 1-year cardiovascular mortality, 30-day moderate to severe PVL, procedure contrast agent volume and procedure time were lower, while transvalvular pressure gradient was higher in BEV. 30-day incidence of stroke, PPI, AKI, major bleeding, MVC and rehospitalization were not statistically different between BEV and SEV. Our study was strengthened by lack of significant heterogeneity across studies and lack of significant difference between RCTs and observational studies for most outcomes.

The higher mortality in SEV than BEV was also robust in currently using new-generation valves, which highlights the importance of our findings. The lack of statistical significance in RCTs may possibly be due to power insufficiency, thus raising the need for an adequately powered RCT comparing different TAVR devices. Consistent with mortality outcome, we also observed a higher incidence of moderate to severe PVL in SEV. Our meta-regression analysis revealed that higher risk for moderate to severe PVL in SEV might partially contribute to its higher mortality, but it needs to be confirmed in future studies due to the exploratory nature of our meta-regression analysis. In agreement with this observation, multiple studies provided relatively convincing evidence that residual moderate to severe PVL was associated with increased short and long-term mortality, similar in BEV and SEV (9, 26). New-generation valves feature sealing skirts or incorporates outer pericardial wrap to minimize PVL (27). Our study and others’ reports confirmed a significant reduction in PVL rates in new-generation valves (28), but the risk difference between BEV and SEV remains substantial and significant. It’s remarkable that <1% of patients developed moderate to severe PVL in patients receiving SAPIEN 3 valves (29).

Conduction abnormalities, including high-grade atrioventricular block leading to PPI, remain the most frequent complication of TAVR (30). The clinical impact of PPI after TAVR remains controversial. Although some studies suggested that periprocedural PPI might be associated with a higher risk of all-cause mortality (31), it’s unlikely to explain the mortality difference observed in our study. The overall incidence of PPI was not statistically different between SEV and BEV. Evolut R and ACURATE neo SEV showed opposite performance regarding PPI risk in our analysis, but both were associated with a higher mortality compared with SAPIEN 3 BEV. In addition, our exploratory meta-regression analysis demonstrated a possible trend toward higher mortality with lower incidence of PPI, which requires further investigation. The adoption of new-generation valves seem to have no evident reduction on PPI incidence (30). On the contrary, SAPIEN 3 valves might even increase PPI risk than its prior-generation valves (29). The low PPI incidence associated with ACURATE neo valve system might be attributed to the relatively low radial force that the inflow portion of this frame exerts on the surrounding conduction system (32).

It is important to note that the potential mortality superiority of BEV to SEV in our analysis was mainly driven from PSM studies but not RCTs. Even by sophisticated matching, these PSM studies were limited by lack of independent adjudication of outcomes and residual confounders that could not be fully accounted for. The choice of TAVR prosthesis strongly depends on the operators’ experience with the devices and patients’ anatomical suitability, both of which were not adequately adjusted in nonrandomized studies (33). Exclusion of certain patients based on anatomic criteria make it impossible to match for such patients’ characteristics in an observational study. In fact, it’s likely that there are considerably more patients with extensive outflow tract calcifications, low implanted coronary arteries, or complex and small femoral access receiving SEV devices. Other confounders not adjusted in PSM studies may include patient risk profiles and TAVR access, etc. Such residual confounders might explain part of the mortality difference between SEV and BEV.

We chose to include PSM studies but not studies with regression analysis and other adjusting methods to pooled with RCTs because the former shares some particular characteristics of RCTs. PSM allows researchers to separate the design from the analysis, and to estimate treatment effects in metrics similar to RCTs (34). While in regression analysis, temptation to model toward a desired result might be present because the outcome is always in sight. It’s simpler and more transparent to assess whether observed confounding has been adequately eliminated in PSM than regression analysis (34, 35). PSM might produce less biased, more robust, and more precise estimates than conventional regression analysis (35, 36), especially when there were few events per confounding variables (37). However, in spite of its advantages, no complete consensus on the comparison of PSM with regression methods has been achieved. With the most appealing transparent advantage of PSM, we were able to evaluate the scientific quality and reporting of PSM studies. In our analysis, all PSM studies described the variables to generate the propensity scores, 9 of 12 studies reported methods of matching algorithm, and 5 described that the matching process was conducted without replacement (Appendix Table 7). Eight studies used the encouraged standardized difference method, which is independent of sample size, as the balance diagnostics to compare the distribution of covariates between matched cohorts. It’s important to note that the 3 PSM studies (9-11) which altogether accounted for more than 80% of the weight in the analysis of the primary outcome, provided appropriate, transparent and detailed descriptions of their statistical methods (Appendix Table 7).

We acknowledged several other limitations in our study. First, the meta-analysis was based on study-level but not patient-level data, and thus we were unable to perform in-depth subgroup analyses or meta-regressions. Second, whether our findings could be generalized to low-risk patients remains unknown. Third, our study included in the SEV group several different TAVR valve systems, however, subgroup analysis showed largely similar results. Fourth, our meta-regression analysis could only be considered as exploratory due to limited number of studies. Fifth, number of trials and events are very small for particular valve types, such as the Portico valve. Sixth, there may be differences in baseline characteristics between subgroups and this might influence the observed outcomes between subgroups, but our analyses revealed no obvious heterogeneity between these subgroups. Seventh, the longest duration of follow-up was limited to 1 year, data on durability and long-term outcomes are needed.

## Conclusions

TAVR with BEV might be associated with a lower early and midterm all-cause and cardiovascular mortality compared with TAVR with SEV. An adequately powered RCT comparing different TAVR devices with longer-term follow-up data are warranted to confirm these findings, and determine their late performance and the effects of underlying risks of patients.

## Data Availability

All data were presented in the main manuscript or supplementary materials.

## Acknowledgements

None.

